# Microbiome Dynamics During Chemoradiotherapy for Anal Cancer

**DOI:** 10.1101/2021.09.07.21263233

**Authors:** Daniel Lin, Molly B. El Alam, Joseph Abi Jaoude, Ramez Kouzy, Jae L. Phan, Jacob H. Elnaggar, Brianna Resendiz, Andrea Y. Delgado Medrano, Erica Lynn, Nicholas D. Nguyen, Sonal S. Noticewala, Geena G. Mathew, Emma B. Holliday, Bruce D. Minsky, Prajnan Das, Van K. Morris, Cathy Eng, Melissa P. Mezzari, Joseph F. Petrosino, Nadim J. Ajami, Ann H. Klopp, Cullen M. Taniguchi, Lauren E. Colbert

## Abstract

**IMPORTANCE:** Patients with localized squamous cell carcinoma of the anus (SCCA) who experience treatment toxicity or recurrences have few therapeutic options.

Investigation into the microbiome’s influence on treatment toxicity or its potential use as a predictive biomarker in this rare disease could improve these patients’ outcomes.

**OBJECTIVE:** To longitudinally characterize the SCCA tumor microbiome and assess its association with treatment-related toxicities.

**DESIGN:** Prospective cohort study.

**SETTING:** Single tertiary cancer center.

**PARTICIPANTS:** Twenty-two patients with biopsy-confirmed non-metastatic SCCA receiving standard-of-care chemoradiotherapy as part of an Institutional Review Board-approved study from April 2017 to July 2019.

**MAIN OUTCOMES AND MEASURES:** Diversity and taxonomic characterization of the SCCA microbiome throughout chemoradiotherapy using swab-based anorectal microbial specimen collection and 16S rRNA gene sequencing.

**RESULTS:** Twenty-two SCCA patients were included in this study with a median (range) age of 58.5 (39-77), and 18 (82%) were women. Alpha diversity remained relatively stable throughout chemoradiotherapy, except for decreases in the Chao1 (*P*=0.03) and Observed Features (*P*=0.03) indices at week 5 relative to baseline. Tumor microbial compositions measured using weighted UniFrac changed significantly by the end of treatment (*P=*0.03). Linear discriminant analysis effect sizes revealed differential enrichment of bacteria at specific time points, including the enrichment of Clostridia at both baseline and follow-up and the enrichment of *Corynebacterium* at week 5. Patients experiencing high toxicity at week 5 had higher relative abundances of Clostridia, Actinobacteria, and Clostridiales at baseline (*P*=0.03 for all).

**CONCLUSIONS AND RELEVANCE:** Our study presents the first longitudinal characterization of the SCCA microbiome throughout chemoradiation. The tumor microbiome undergoes significant changes during and after chemoradiotherapy, and patient-reported toxicity levels are associated with patients’ microbial profiles. Further studies into these microbial characterizations and associations are needed to elucidate the tumor microbiome’s role in predicting treatment-related outcomes for SCCA patients.

**Key Points:** *QUESTION:* How does the squamous cell anal tumor microbiome change during chemoradiotherapy, and how do these changes influence treatment-related toxicity?

*FINDINGS:* Prospective longitudinal characterization of anal cancer patients revealed significant modulation of the local tumor microbiome in response to chemoradiotherapy including shifts in overall composition and differential enrichment of key taxa. Additionally, enrichment of specific taxa at baseline was associated with increased levels of treatment-related toxicities by the end of treatment.

*MEANING:* The anal tumor microbiome is significantly altered by chemoradiotherapy and could potentially serve as a biomarker for treatment-related toxicities.

## Introduction

Squamous cell carcinoma of the anus (SCCA) is a rare gastrointestinal cancer that is linked to prior human papillomavirus (HPV) infections in approximately 90% of cases.^1^ The standard-of-care treatment for patients with localized SCCA is chemoradiotherapy (CRT) which leads to 5-year survival rates of 75%.^2^ However, 30-40% of patients with advanced SCCA experience local recurrence or treatment-related toxicity, for which there are no effective therapeutic options.^3–7^ Despite the use of HPV vaccines, the incidence and morbidity from SCCA continue to rise.^8^

The human microbiome influences the oncogenesis and treatment response of many cancers, including gastrointestinal cancers,^9–11^ through the modulation of inflammation, cell proliferation, and metabolism.^12, 13^ Clinical and preclinical data suggest that gut microbiota are direct modulators of therapy response and toxicity.^14–18^

Previous studies have delineated the microbiomes of HPV-driven cancers of the cervix and oropharynx, but the role of microbiota in SCCA remains unknown.^19–21^ Studies in cervical cancer revealed significant associations between the gut microbiome and treatment-related toxicity.^21, 22^ However, little is known about the microbiome’s influence on treatment in SCCA and whether it can be used as a predictor of patient-reported toxicity. Therefore, we sought to longitudinally characterize the tumor microbiome and its relationship to acute gastrointestinal toxicity in SCCA patients undergoing modern CRT.

## Methods

### Patients

Anorectal tumor swabs were prospectively collected from SCCA patients receiving standard-of-care treatment as part of an Institutional Review Board–approved study (protocol #2014-0543) at M.D. Anderson Cancer Center from April 2017 to July 2019. Patients with biopsy-confirmed non-metastatic SCCA with a palpable tumor were included in the study. Patients with a history of abdominal or pelvic radiotherapy were excluded. Written informed consent was obtained from all patients. Eligible patients received intensity-modulated radiotherapy (IMRT), volumetric modulated arc therapy (VMAT), or proton therapy with concurrent chemotherapy. All patients received a minimum radiation dose of 46 Gy in 23 fractions over 5 weeks. Patients’ demographic data were collected from medical records.

### Patient-Reported Outcomes and Adverse Events

Patients completed the bowel subdomain of the Expanded Prostate Cancer Index Composite (EPIC) questionnaire before (baseline), during (weeks 1, 3, and 5), and after treatment (week 12 [follow-up]) as described previously.^23^ The EPIC bowel subdomain is divided into “Function” and “Bother” subdomains, each comprising of 7 questions. To highlight the differences in microbial composition at each time point, we calculated a patient-reported outcome score for each patient by comparing the patient’s EPIC score to the median. Patients with EPIC scores greater than the median had low toxicity, and those with lower scores had high toxicity. At the same time points, a physician assigned each patient a Common Terminology Criteria for Adverse Events (CTCAE; version 5.0) score for anal dermatitis, and patients were stratified based on scores of 0-1 and 2-3.

### 16S rRNA Gene Sequencing

Anorectal swab samples were collected immediately before (baseline), during (weeks 1, 3, and 5), and after treatment (week 12 [follow-up]) (**Figure 1A**). Week 1 samples were collected after patients had received at minimum 1 fraction of radiotherapy. A digital rectal exam was performed with a sterile gloved finger to palpate the anal tumor, after which an Isohelix DNA Buccal Swab (Isohelix, SK-2S) was used to collect the tissue and fecal material. Samples were stabilized using BuccalFix DNA Stabilization solution (Isohelix, BFX-25) within 1 hour of collection and stored at −80°C until DNA isolation was performed using the MoBIO PowerSoil kit (Qiagen, 12855-50).

**Figure 1.**
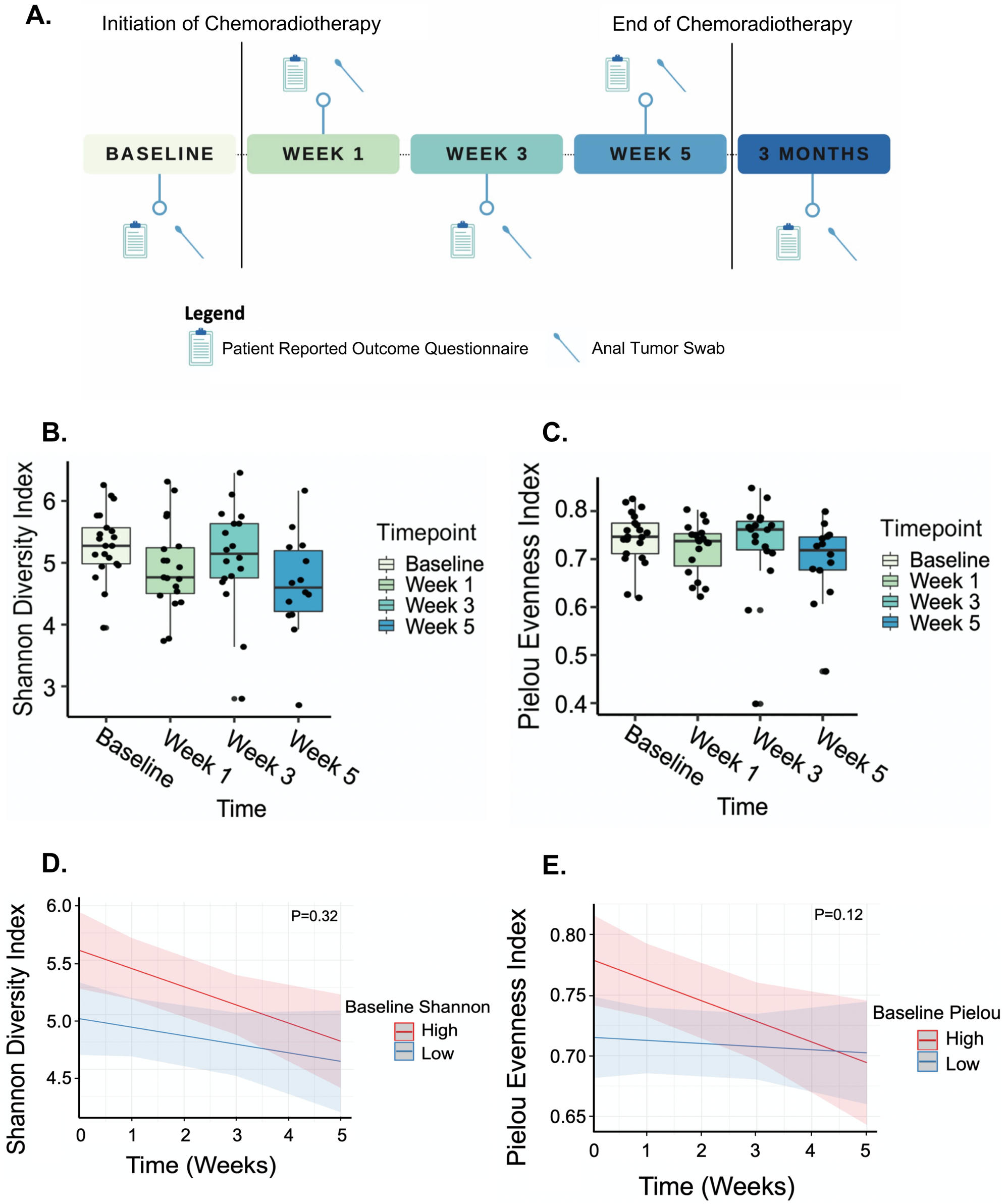
Alpha Diversity During Chemoradiotherapy. **A,** Study overview depicting the time points at which anorectal swabs were collected and patient-reported outcome questionnaires were administered. **B and C,** SCCA patients’ Pielou evenness (B) and Shannon diversity (C) indices did not change significantly throughout chemoradiotherapy. Wilcoxon signed-rank test was used for comparisons. **D and E,** Linear mixed modeling revealed trends of high and low baseline alpha diversity metrics predicting diversity richness (D) and evenness (E).

The 16S rRNA gene amplification and sequencing methods were adapted from the Human Microbiome Project and Earth Microbiome Project.^24^ The V4 region of the 16S rRNA marker gene was PCR-amplified using a 515F-806R primer pair. The Alkek Center for Metagenomics and Microbiome Research at Baylor College of Medicine performed the 16S rRNA gene sequencing using the Illumina MiSeq platform and a 2×250bp paired-end protocol.

### Microbiome Sequencing Analysis

To analyze the 16S rRNA gene reads, we utilized the QIIME2 microbiome bioinformatics platform (v2019.10).^25^ A sampling depth of 9160 sequences per sample was used for downstream comparative analysis. Sequence quality control and amplicon sequence variant feature table construction were performed using denoising via DADA2.^26^ Phylogenetic reference tree construction was performed using the SILVA 128 database, which served as the reference tree backbone for fragment insertion.^27^ Taxonomic identification was assigned using a naïve Bayes classifier trained using the SILVA 132 database.

### Microbial Diversity Metrics

We analyzed the 16S rRNA sequencing data using 7 different alpha diversity metrics. The Shannon diversity index accounts for richness and evenness of taxa within a sample. The inverse Simpson diversity index measures the relative abundance of species that make up the richness.^28, 29^ The Chao1 diversity index accounts for the number of rare taxa likely missed owing to undersampling.^30^ Faith phylogenetic diversity (PD) accounts for the phylogenetic differences between species in a sample.^31^ The Fisher index quantifies the relationship between the number and abundance of each species in a sample.^32^ Pielou evenness index calculates the proportions of individual species within a sample population.^33^ Finally, Observed Features provides a count of species with at least one read.^34^

### Microbial Composition

Linear discriminant analysis (LDA) effect size (LEfSe) was used to identify bacterial taxa that were differentially enriched in samples at baseline as compared to week 5 and at week 5 as compared to follow-up.^35^ An LDA score cut-off of 4.0 was used. The alpha value for the Kruskal-Wallis test among classes was set at 0.05. Weighted and unweighted UniFrac distances and Bray-Curtis dissimilarity were used to generate coordinates for each sample. Principal coordinate analysis was used to visualize similarities and dissimilarities between time point groups. Distances between coordinate groups were compared using permutational multivariate analysis of variance (PERMANOVA). We used LefSe with an LDA score cut-off of 4.0 to assess the differences in the baseline microbial profiles of patients with low and high toxicity at the end of treatment.

### Statistical Analysis

We used descriptive statistics to summarize patients’ demographic and clinical characteristics. To assess changes in microbiome diversities, we used Wilcoxon signed-rank tests to compare the alpha diversity indices at weeks 1, 3, and 5 and follow-up to those at baseline. We built a linear mixed model using time as the independent predictor of diversity to include patients who did not have samples available at all time points (n=14). We used individual patients as a random effect and an interaction term of baseline diversity and time as the main covariate. We used a Wilcoxon signed-rank test to detect significant differences in the relative abundances of taxa identified using LEfSe throughout treatment.

We compared the alpha diversity indices between patients with high or low toxicity using a Wilcoxon rank-sum test. We assessed the differences in identified taxa between the two toxicity groups using a Wilcoxon rank-sum test.

We compared patients’ clinical and demographic characteristics according to their CTCAE scores and EPIC scores. Finally, we tested for associations between baseline alpha diversity and clinical characteristics using the Kruskal-Wallis test and Spearman correlation. Statistical significance was set at an alpha of 5% for a two-sided *P* value. All analyses were performed using QIIME2 (v2019.10) and Rstudio Orange Blossom.

## Results

### Patient Characteristics

The study included 22 patients (18 women [82%] and 4 men [18%]), whose median age was 58.5 years (range, 39-77 years). Patients’ baseline characteristics are shown in **Table 1**. Most patients received chemotherapy with cisplatin and 5-FU (18 [86%]) and underwent IMRT or VMAT (19 [91%]). Patients received a median radiation dose of 54 Gy over a median of 27 fractions. During the study, 7 patients (33%) received antibiotics. Baseline swab samples were collected from 21 patients; week 1 samples, from 19 patients; week 3 samples, from 18 patients; week 5 samples, from 14 patients; and follow-up samples, from 9 patients (**Supplemental Table 1**). Patients’ clinical characteristics were not significantly associated with either toxicity measurement. Additionally, baseline alpha diversity was not significantly associated with any patient characteristics.

**Table 1.**
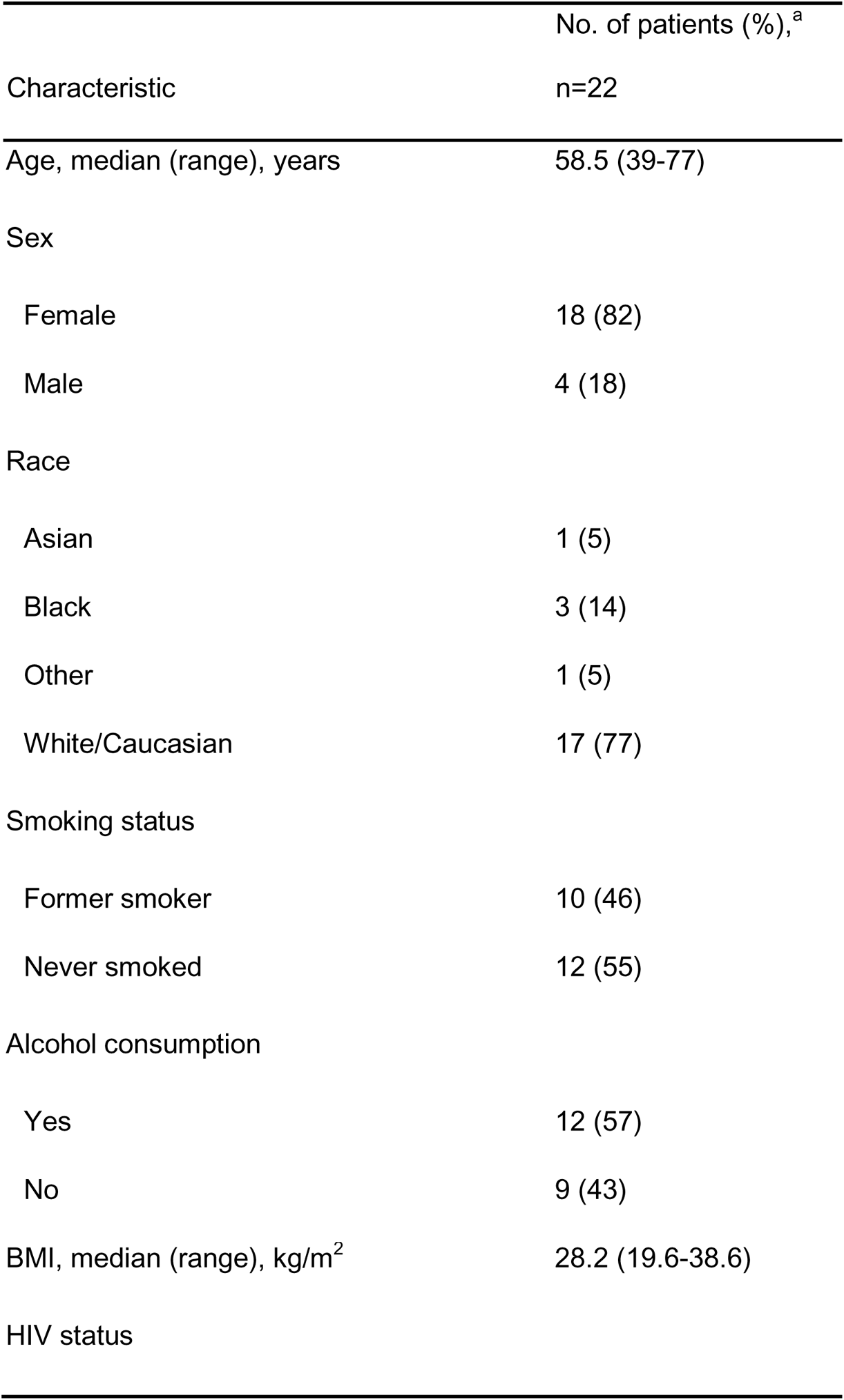
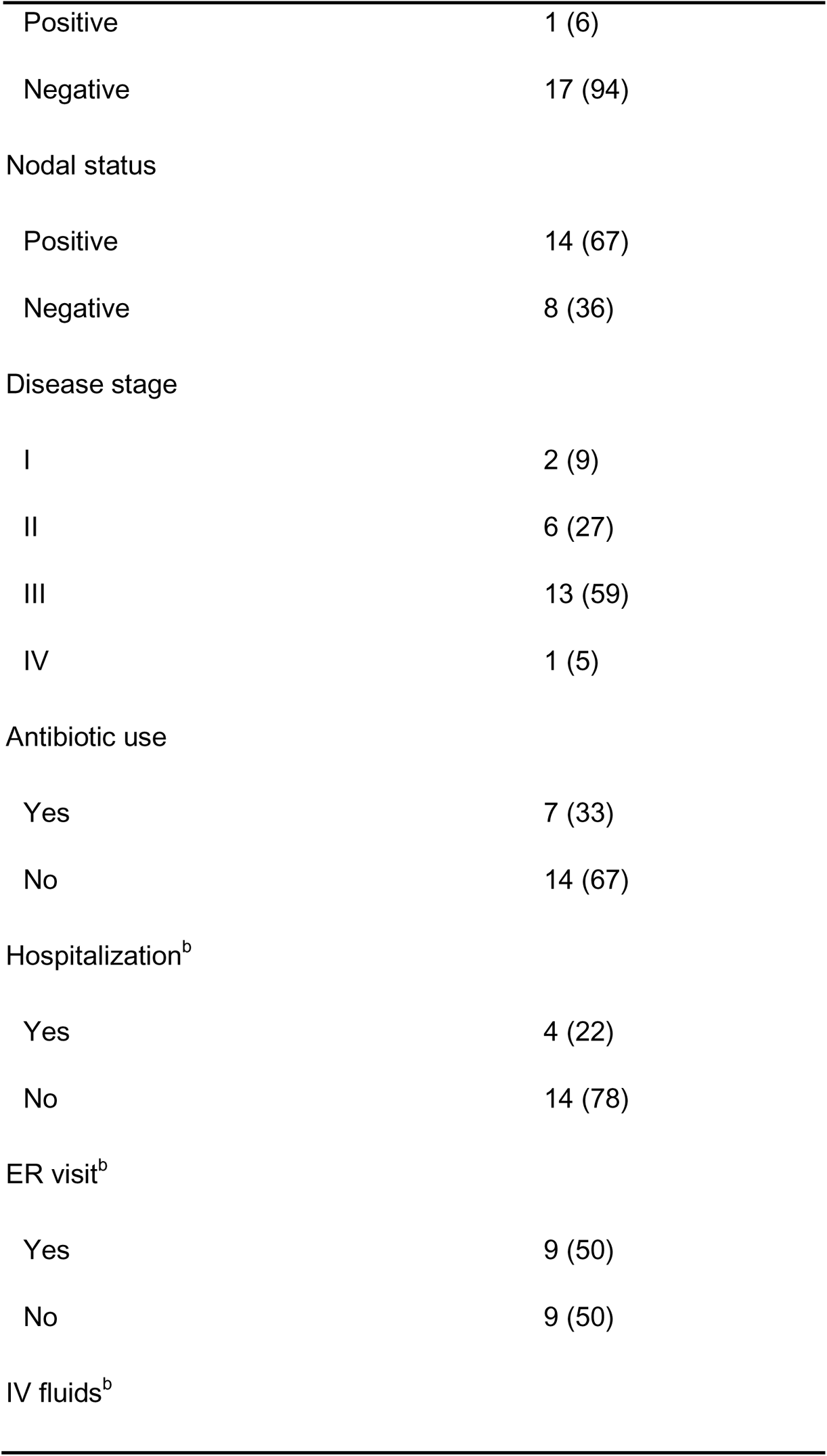
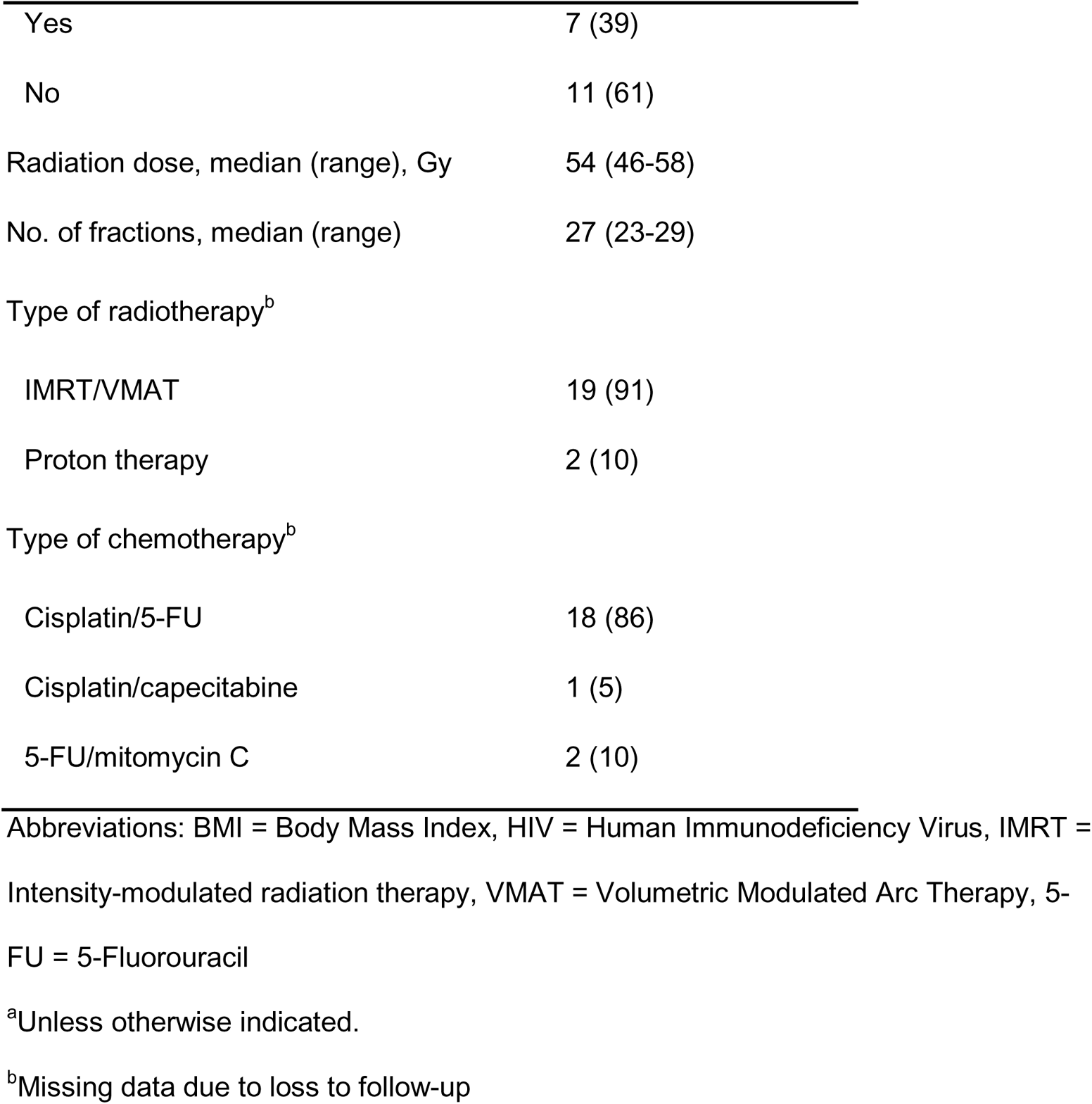
Patients’ Demographic and Clinical Characteristics.

|  | No. of patients (%),<sup>a</sup> |
| --- | --- |
| Characteristic | n=22 |
| Age, median (range), years | 58.5 (39-77) |
| Sex |  |
| Female | 18 (82) |
| Male | 4 (18) |
| Race |  |
| Asian | 1 (5) |
| Black | 3 (14) |
| Other | 1 (5) |
| White/Caucasian | 17 (77) |
| Smoking status |  |
| Former smoker | 10 (46) |
| Never smoked | 12 (55) |
| Alcohol consumption |  |
| Yes | 12 (57) |
| No | 9 (43) |
| BMI, median (range), kg/m<sup>2</sup> | 28.2 (19.6-38.6) |
| HIV status |  |
| Positive | 1 (6) |
| Negative | 17 (94) |
| Nodal status |  |
| Positive | 14 (67) |
| Negative | 8 (36) |
| Disease stage |  |
| I | 2 (9) |
| II | 6 (27) |
| III | 13 (59) |
| IV | 1 (5) |
| Antibiotic use |  |
| Yes | 7 (33) |
| No | 14 (67) |
| Hospitalization<sup>b</sup> |  |
| Yes | 4 (22) |
| No | 14 (78) |
| ER visit<sup>b</sup> |  |
| Yes | 9 (50) |
| No | 9 (50) |
| IV fluids<sup>b</sup> |  |
| Yes | 7 (39) |
| No | 11 (61) |
| Radiation dose, median (range), Gy | 54 (46-58) |
| No. of fractions, median (range) | 27 (23-29) |
| Type of radiotherapy<sup>b</sup> |  |
| IMRT/VMAT | 19 (91) |
| Proton therapy | 2 (10) |
| Type of chemotherapy<sup>b</sup> |  |
| Cisplatin/5-FU | 18 (86) |
| Cisplatin/capecitabine | 1 (5) |
| 5-FU/mitomycin C | 2 (10) |

Abbreviations: BMI = Body Mass Index, HIV = Human Immunodeficiency Virus, IMRT = Intensity-modulated radiation therapy, VMAT = Volumetric Modulated Arc Therapy, 5-FU = 5-Fluorouracil
<sup>a</sup>Unless otherwise indicated.
<sup>b</sup>Missing data due to loss to follow-up

### Tumor Microbiome Characteristics

Alpha diversity measurements remained relatively stable throughout treatment, with the Shannon, Pielou, Simpson, inverse Simpson, and Faith PD indices showing no significant changes in community richness or evenness; however, Observed Features and Chao1 indices revealed significant decreases as compared to baseline (**Figure 1B,C**, **Supplemental Table 2**). Linear mixed modeling using time as a predictor of decreased diversity revealed similar trends. Shannon and Pielou indices did not show a significant association with time as a predictor (**Figure 1D,E**). However, when accounting for high or low baseline diversity, patients with higher baseline Observed Features, Chao1 diversity, and Faith PD indices tended to have faster declines in diversity (*P*<0.05; **Supplemental Table 3**).

Beta diversity analysis, accounting for differences in the abundance of taxa and phylogenetic relatedness, demonstrated significant shifts in the overall community composition from baseline to week 5. The weighted UniFrac distance matrices generated from baseline and week 5 samples are illustrated in **Figure 2A**. A comparison of these distance coordinates revealed significant changes in the overall microbial compositions at week 5 as compared to baseline (*P=*0.032). Comparisons of the baseline and week 5 samples using unweighted UniFrac distances and Bray-Curtis index yielded similar results (**Supplemental Table 4**).

**Figure 2.**
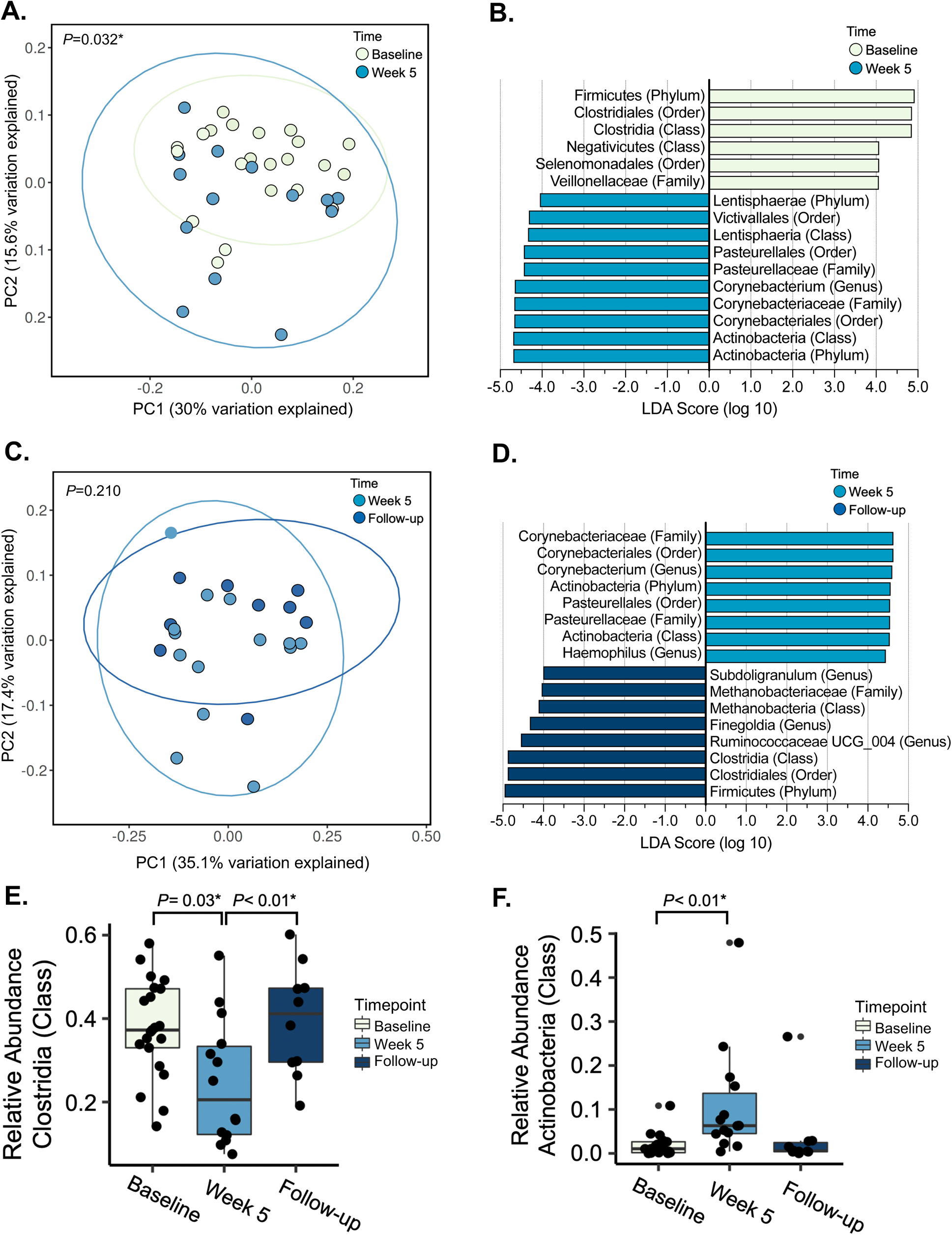
Beta Diversity and Taxa-Specific Changes During Chemoradiotherapy. **A,** Principal coordinate analysis was used to visualize microbial composition of samples collected at baseline and week 5 using weighted UniFrac distances. PERMANOVA revealed significantly different microbial compositions at baseline and week 5. **B,** LEfSe revealed time point–specific taxa enrichment at baseline as compared to week 5 (LDA threshold, 4.0). The top 3 taxa enriched at baseline were hierarchically related, as were the top 5 taxa enriched at week 5. **C,** Principal coordinate analysis was used to visualize microbial composition of samples collected at week 5 and follow-up using weighted UniFrac distances. PERMANOVA showed no significant differences in patients’ microbiome profiles at week 5 as compared to follow-up. **D,** LEfSe revealed time point–specific taxa enrichment at week 5 as compared to follow-up (LDA threshold, 4.0). The taxa enriched at week 5 as compared to follow-up were strikingly similar to the taxa enriched at week 5 as compared to baseline, with 7 overlapping taxa. In addition, the top 3 taxa enriched at follow-up as compared to week 5 were the same as the top 3 taxa enriched at baseline as compared to week 5. **E and F,** The relative abundances of Clostridia (E) and Actinobacteria (F) change significantly throughout chemoradiotherapy. The abundance of Clostridia decreases significantly from baseline to week 5 but returns to its baseline level by follow-up. In contrast, the abundance of Actinobacteria increases significantly from baseline to week 5 and then decreases (though not significantly) from week 5 to follow-up. Wilcoxon signed-rank test was used for comparisons.

LEfSe identified 16 taxa that were differentially enriched at baseline (6 taxa) or week 5 (10 taxa) (**Figure 2B**). The relative abundances of 11 of these taxa significantly differed between baseline and week 5 (**Supplemental Table 5**). Upon closer inspection, the top 3 taxa enriched at baseline (Firmicutes [phylum], Clostridia [class], and Clostridiales [order]) were hierarchically related. Similarly, the top 5 taxa enriched at week 5 (Actinobacteria [phylum], Actinobacteria [class], Corynebacteriales [order], *Corynebacteriaceae* [family], and *Corynebacterium* [genus]) were also related. The weighted UniFrac profiles of the week 5 and follow-up samples did not differ significantly (*P*=0.210) (**Figure 2C**). LEfSe revealed that some taxa enriched at follow-up were the same as those previously enriched at baseline when both were compared with week 5. Follow-up samples had greater abundances of Firmicutes (phylum), Clostridiales (order), and Clostridia (class) when compared with week 5 samples (**Figure 2D**). Furthermore, the top 7 taxa enriched at week 5 as compared with follow-up were identical to the top 7 taxa enriched at week 5 when compared with baseline.

The relative abundances of Clostridia (class) and Clostridales (order) decreased during CRT (*P*=0.033 for both) but increased by follow-up (*P*=0.004 for both) (**Figure 2E**, **Supplemental Table 5**). Similarly, the relative abundances of Corynebacteriales (order), *Corynebacteriaceae* (family), and *Corynebacterium* (genus) increased during treatment (*P*=0.002, 0.002, and 0.001, respectively) but returned to baseline levels by follow-up (*P*=0.002, 0.002, and 0.006, respectively). The relative abundances of *Finegoldia* (genus) and *Subdoligranulum* (genus) did not change during treatment but increased following treatment (*P*=0.033 and 0.006, respectively). The relative abundance of Actinobacteria (class) increased during treatment (*P*=0.003) but did not change after treatment (**Figure 2F**). These findings indicate that although the composition of the microbiome changes during treatment, there is a period of microbial restoration following treatment during which the local anal microbiome begins to resemble the baseline microbiome.

### CTCAE Scores and Patient-Reported Toxicity

16 patients had CTCAE scores of 2-4, and 3 had scores of 0-1. Of the 12 patients who answered the EPIC questionnaire, 6 had low scores and 6 had high scores. Baseline alpha diversity did not differ significantly between patients with CTCAE scores of 2-4 or 0-1 at week 5 (**Figure 3A**), except that patients with higher CTCAE scores had a significantly higher median Pielou index at baseline (0.75 vs. 0.69; *P*=0.02) (**Figure 3B**). Baseline alpha diversity did not differ significantly between patients with low or high EPIC scores (**Figure 3C, D**).

**Figure 3.**
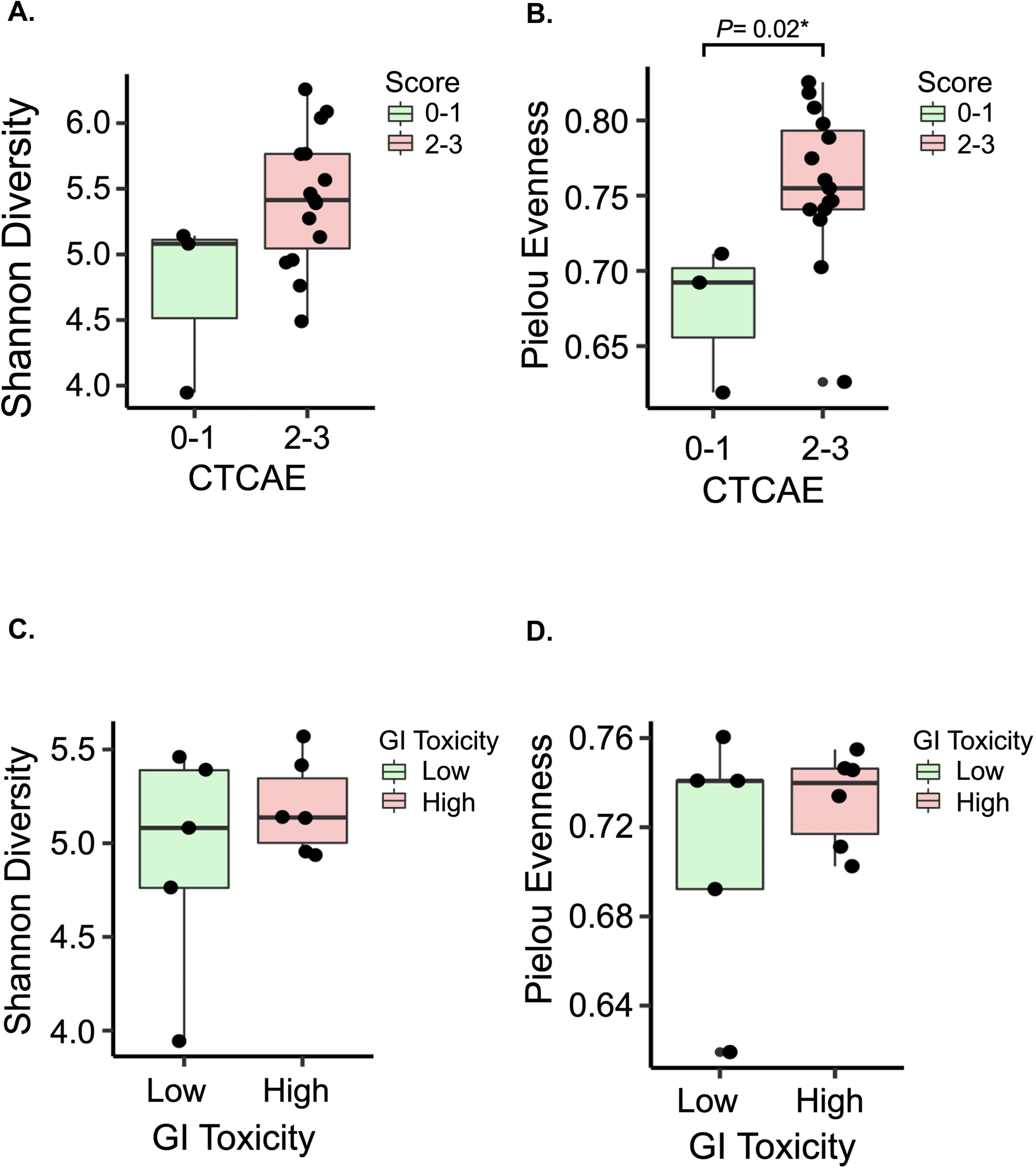
Analysis of Alpha Diversity Metrics and Quality-of-Life Measurements. **A and B,** Increased microbial richness (A) and evenness (B) at baseline were associated with higher CTCAE scores at week 5. Wilcoxon rank-sum test was used for comparisons. **C and D,** Microbial richness (A) and evenness (B) at baseline were not associated with toxicity at week 5. Wilcoxon rank-sum test was used for comparisons.

LEfSe identified several differences in the baseline abundances of species in samples from patients with low or high toxicity at week 5. Patients with low toxicity at week 5 had higher levels of Sellimonas, whereas patients with high toxicity at week 5 had higher levels of Actinobacteria (phylum and class), Peptoniphilus, Clostridiales, Clostridia, and *Clostridiales FamilyXI* (**Figure 4A**). Patients with high toxicity at week 5 had significantly higher relative abundances of Clostridia, Actinobacteria (class), and Clostridiales at baseline (*P*=0.03 for all), whereas patients with low toxicity had a significantly higher relative abundance of *Sellimonas* (genus; *P*=0.048) (**Figure 4B,C**, **Supplemental Table 6**).

**Figure 4.**
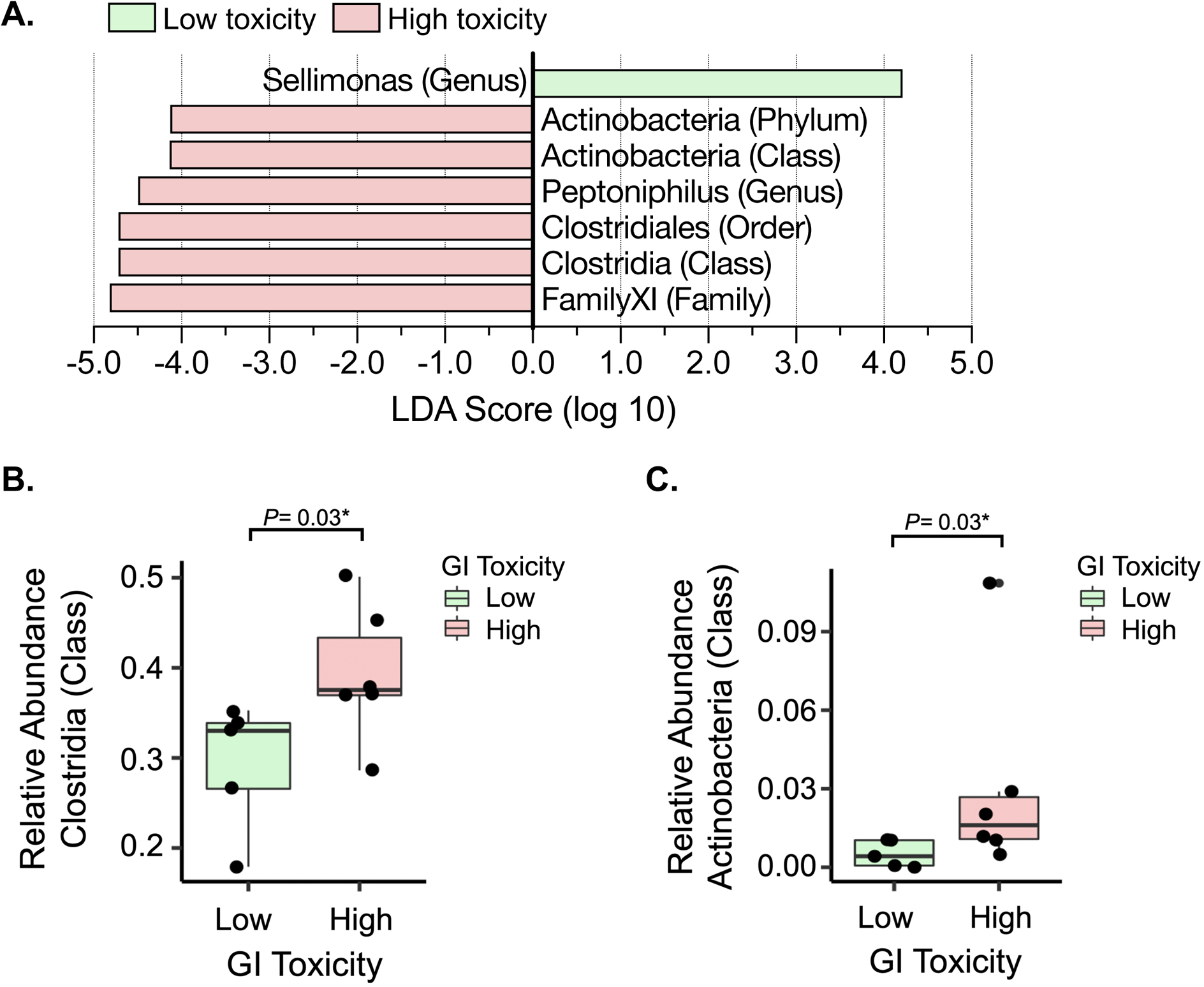
Baseline Taxonomic Enrichment May Serve as a Biomarker for Chemoradiotherapy Quality of Life at Week 5. **A,** LEfSe of baseline microbial profiles revealed differences in taxa-specific enrichment between patients grouped by low and high toxicity based on EPIC scores. **B and C,** The relative abundances of Clostridia (B) and Actinobacteria (C) at baseline differed significantly between patients grouped by high and low toxicity based on EPIC scores. Wilcoxon rank-sum test was used for comparisons.

## Discussion

While the microbiome has been well characterized in HPV-driven cancers, published research describing the SCCA microbiome remains scarce.^36^ Here, we present the first longitudinal characterization of the SCCA microbiome and its response to CRT. Alpha diversity metrics revealed relative stability within the local anal tumor microbiome throughout treatment. However, we observed a trend of decreasing alpha diversity during treatment when accounting for baseline diversity. We suspect that, with increasing patient numbers, this trend of decreasing diversity during CRT would become more prominent, as described in cervical cancer patients.^37^

In our cohort, beta diversity metrics indicated a distinct shift in the overall composition of the microbiome during CRT. Further investigation using LEfSe and relative abundance comparisons revealed 11 taxa, including Clostridiales, Corynebacteriales, and Actinobacteria (class), that were differentially enriched by the end of CRT. Many of these enriched taxa directly overlapped with the LEfSe results comparing week 5 samples to follow-up where samples exhibited similar taxa enriched at baseline. These findings suggested a period of restoration of the anal microbiome in the weeks following CRT.

Our quantitative data also offer new insight into the microbiome of SCCA patients and its association with their quality of life. Our study prospectively measured toxicity using both patient and physician assessment tools to help capture the true extent of disease- and treatment-related toxicity. Microbial diversity did not predict toxicity, but some bacterial species were significantly associated with CRT-related toxicity. LEfSe and relative abundance comparisons revealed that higher relative abundances of bacterial taxa such as Clostridia and Actinobacteria (class) at baseline were associated with increased toxicity at the end of treatment. Additional studies are needed to determine whether certain microbial species lead to increased gastrointestinal toxicity or vice-versa.

Our study suggests that CRT affects the overall composition of the SCCA microbiome and that specific microbes in the local tumor microenvironment may play a role in treatment-related toxicity. However, the mechanisms of such interactions undetermined. One such mechanism may involve the local breakdown of the epithelial barrier due to radiotherapy. The reduced epithelial integrity, change in mucosal secretions, and damage to mucosal immune cells could enable the translocation and persistence of abnormal microbiota.^38^ The outgrowth of certain bacterial species on the epithelial membrane due to CRT could affect gastrointestinal and skin toxicity through inflammation and immune regulation. Furthermore, following radiotherapy, the epithelial lining can begin healing, which could allow for the return of bacteria native to the mucosa.^39, 40^ In addition, preclinical studies suggest that the microbiome influences radiation proctitis, resulting in the modulation of cytokines, particularly interleukin 1β.^41^ The immune modulation of increased interleukin 1β has been similarly observed in bacterial vaginosis, which is associated with HPV-driven cervical cancer.^42^ Lastly, radiotherapy-induced diarrhea, a common toxicity of pelvic radiotherapy, has been linked to significant shifts in the gut microbiome, with some studies suggesting that gut dysbiosis is a predictor of severe diarrhea during and after radiotherapy.^43, 44^ These possible mechanisms require further investigation in both preclinical and clinical studies.

Multiple studies have investigated changes in microbiome profiles during pelvic radiation, but none have characterized the SCCA microbiome.^43–45^ Previous clinical studies have demonstrated a similar marked decrease in Firmicutes in the gut resulting from radiotherapy.^43, 46^ Also, preclinical studies of radiation-induced rectal dysbiosis showed similar decreases in Clostridia, Clostridiales, and Firmicutes.^41, 43^ We observed that an increased baseline level of Clostridiales was associated with higher toxicity during treatment, which is contrary to previous reports of the potential positive effects of Clostridiales during CRT for HPV-driven cervical cancer.^22, 37^

One of the most significantly enriched bacteria in our end-of-treatment samples was *Corynebacterium*. *Corynebacterium* is believed to be largely commensal inhabitants of human skin flora, but its role in the tumor microbiome and treatment response is unclear. However, some members of *Corynebacterium,* such as *Corynebacterium jeikeium,* may prevent oxidative damage to both itself and epidermal tissue through its acquisition of manganese necessary for the function of manganese-dependent superoxide dismutase.^47^

The present study had some limitations. First, it was a single-institution study with a limited sample size. Thus, its results may not be generalizable to a broader population considering the known regional variations in microbial communities.^48^ However, given the low prevalence of SCCA, this study was the first and largest longitudinal study to analyze this population’s microbiome during CRT. Second, the standard-of-care chemotherapy at our institution is cisplatin and 5-FU, which might have different toxicity and microbiome interactions than the widely used regimen of 5-FU and mitomycin-C. However, since cisplatin is the most commonly used radio-sensitizing chemotherapy for HPV-related malignancies, these data may be valuable in comparing microbiome effects between other anatomical sites such as oropharyngeal cancer or cervical cancer. Third, our study focused on acute changes occurring within a relatively short follow-up time (3 months). Additionally, half the patients did not provide patient-reported outcome data at the final follow-up. The patient attrition rate might be due to the increase in loss to follow-up after the treatment was concluded at week 5. However, longitudinal sampling is one of the most robust methods to evaluate microbiome data. In addition, we used appropriate statistical analyses to account for the small sample size. Finally, in-depth research involving preclinical models could provide unique insights into the role of the microbiota in this disease, but the HPV-driven tumor and the human microbiome remain difficult to recapitulate in animal models. Our hypothesis-generating work is the first to shed light on the microbiome’s role in SCCA and its associations with toxicity, but future studies will be essential to confirm and expand upon these findings.

In conclusion, we found that the local tumor microbiome of SCCA patients changes significantly throughout and following CRT. Our findings suggest an association between the microbial profiles and toxicity levels of SCCA patients receiving CRT. Our work highlights the microbiome’s associations and potential roles in an understudied disease and underreported quality-of-life issues. Our data help provide a foundation for future research to better understand the potentially predictive and therapeutic role of microbiota in SCCA.

## Data Availability

Dr. Cullen M. Taniguchi and Dr. Lauren E. Colbert had complete access to the data in the study and take responsibility for the integrity and accuracy of the data analysis.

## Author Contributions

Dr. Cullen Taniguchi and Dr. Lauren Colbert had complete access to the data in the study and take responsibility for the integrity and accuracy of the data analysis.

## Concept and design

Colbert, Taniguchi, Klopp

## Acquisition, analysis, or interpretation of data

Lin, El Alam, Abi Jaoude, Kouzy, Phan, Delgado Medrano, Lynn, Elnaggar, Mathew, Holliday, Mezzari, Petrosino, Ajami, Klopp, Colbert, Taniguchi

## Drafting of the manuscript

Lin, El Alam, Kouzy, Abi Jaoude, Elnaggar, Resendiz, Klopp, Colbert, Taniguchi

## Critical revision of the manuscript for important intellectual content

Lin, El Alam, Abi Jaoude, Kouzy, Phan, Elnaggar, Resendiz, Delgado Medrano, Lynn, Nguyen, Noticewala, Mathew, Holliday, Minsky, Das, Morris, Eng, Mezzari, Petrosino, Ajami, Klopp, Colbert, Taniguchi

## Statistical analysis

El Alam, Abi Jaoude, Kouzy, Lin

## Obtained funding

Taniguchi, Colbert, Klopp

## Administrative, technical, or material support

Noticewala, Mathew, Holliday, Minsky, Das, Morris, Eng, Mezzari, Petrosino, Ajami

## Supervision

Taniguchi, Colbert

## Additional Contributions

We thank the patients for their immeasurable contribution and the radiation oncology clinical and lab teams at the University of Texas MD Anderson Cancer Center.

---

**Supplemental Table 1.** Sample Availability for Each Patient at Each Time Point.

| Patient | Baseline | Week 1 | Week 3 | Week 5 | Follow-up |
| --- | --- | --- | --- | --- | --- |
| 1 | Present | Present | Present | Present | Present |
| 2 | Present | Present | Present | Present | Present |
| 3 | Present | Present | Present | Present | Present |
| 4 | Present | Present | Present | Present | Present |
| 5 | Present | Present | Present | **Absent** | **Absent** |
| 6 | Present | Present | Present | **Absent** | **Absent** |
| 7 | Present | Present | **Absent** | **Absent** | **Absent** |
| 8 | Present | Present | Present | Present | Present |
| 9 | Present | Present | Present | Present | **Absent** |
| 10 | Present | Present | Present | Present | **Absent** |
| 11 | Present | Present | Present | Present | Present |
| 12 | Present | Present | Present | Present | Present |
| 13 | Present | Present | Present | **Absent** | **Absent** |
| 14 | Present | Present | Present | Present | Present |
| 15 | **Absent** | Present | **Absent** | Present | Present |
| 16 | Present | Present | Present | **Absent** | **Absent** |
| 17 | Present | **Absent** | Present | Present | **Absent** |
| 18 | Present | **Absent** | Present | **Absent** | **Absent** |
| 19 | Present | ***Absent*** | ***Absent*** | ***Absent*** | ***Absent*** |
| 20 | Present | Present | ***Absent*** | ***Absent*** | ***Absent*** |
| 21 | Present | Present | Present | Present | ***Absent*** |
| 22 | Present | Present | Present | Present | ***Absent*** |

**Supplemental Table 2.**
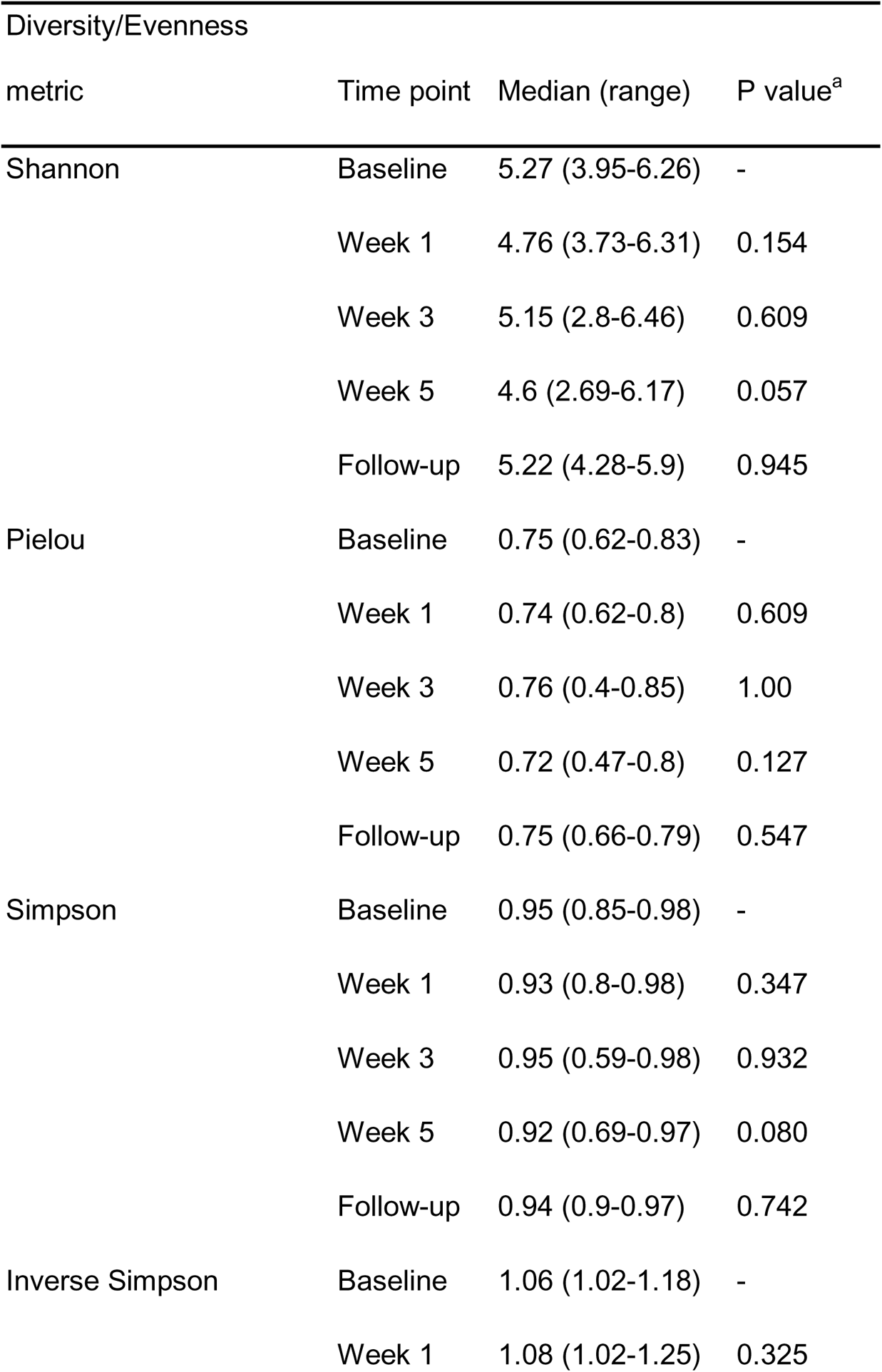
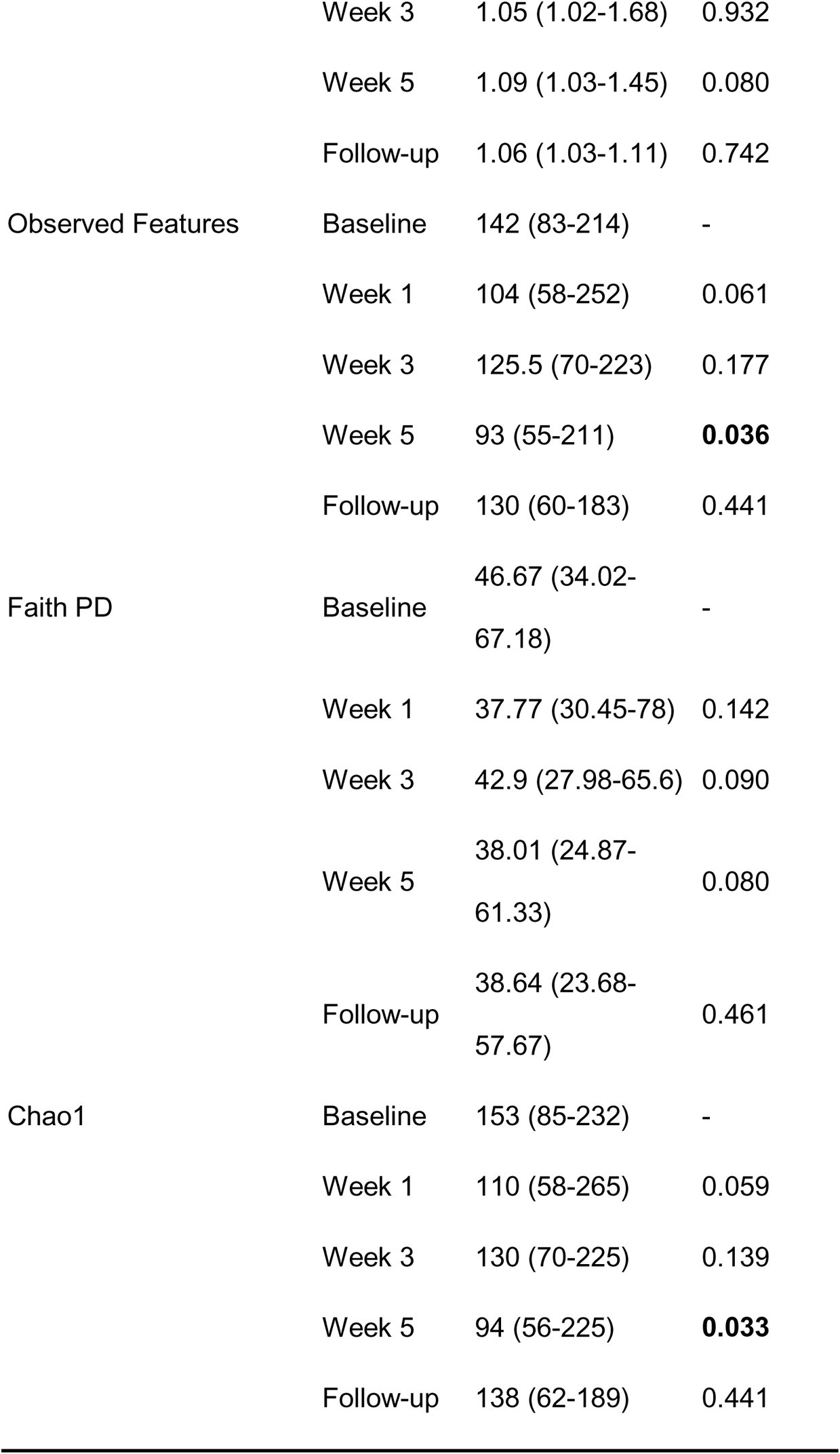
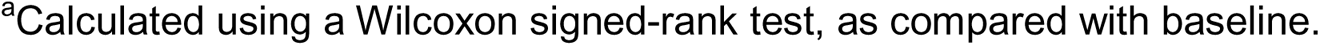
Diversity and Evenness Metrics at Baseline vs. Those During and After Chemoradiotherapy.

| Diversity/Evenness |  |  |  |
| --- | --- | --- | --- |
| metric | Time point | Median (range) | P value<sup>a</sup> |
| Shannon | Baseline | 5.27 (3.95-6.26) | - |
|  | Week 1 | 4.76 (3.73-6.31) | 0.154 |
|  | Week 3 | 5.15 (2.8-6.46) | 0.609 |
|  | Week 5 | 4.6 (2.69-6.17) | 0.057 |
|  | Follow-up | 5.22 (4.28-5.9) | 0.945 |
| Pielou | Baseline | 0.75 (0.62-0.83) | - |
|  | Week 1 | 0.74 (0.62-0.8) | 0.609 |
|  | Week 3 | 0.76 (0.4-0.85) | 1.00 |
|  | Week 5 | 0.72 (0.47-0.8) | 0.127 |
|  | Follow-up | 0.75 (0.66-0.79) | 0.547 |
| Simpson | Baseline | 0.95 (0.85-0.98) | - |
|  | Week 1 | 0.93 (0.8-0.98) | 0.347 |
|  | Week 3 | 0.95 (0.59-0.98) | 0.932 |
|  | Week 5 | 0.92 (0.69-0.97) | 0.080 |
|  | Follow-up | 0.94 (0.9-0.97) | 0.742 |
| Inverse Simpson | Baseline | 1.06 (1.02-1.18) | - |
|  | Week 1 | 1.08 (1.02-1.25) | 0.325 |
|  | Week 3 | 1.05 (1.02-1.68) | 0.932 |
|  | Week 5 | 1.09 (1.03-1.45) | 0.080 |
|  | Follow-up | 1.06 (1.03-1.11) | 0.742 |
| Observed Features | Baseline | 142 (83-214) | - |
|  | Week 1 | 104 (58-252) | 0.061 |
|  | Week 3 | 125.5 (70-223) | 0.177 |
|  | Week 5 | 93 (55-211) | **0.036** |
|  | Follow-up | 130 (60-183) | 0.441 |
| Faith PD | Baseline | 46.67 (34.02-67.18) | - |
|  | Week 1 | 37.77 (30.45-78) | 0.142 |
|  | Week 3 | 42.9 (27.98-65.6) | 0.090 |
|  | Week 5 | 38.01 (24.87-61.33) | 0.080 |
|  | Follow-up | 38.64 (23.68-57.67) | 0.461 |
| Chao1 | Baseline | 153 (85-232) | - |
|  | Week 1 | 110 (58-265) | 0.059 |
|  | Week 3 | 130 (70-225) | 0.139 |
|  | Week 5 | 94 (56-225) | **0.033** |
|  | Follow-up | 138 (62-189) | 0.441 |

<sup>a</sup>Calculated using a Wilcoxon signed-rank test, as compared with baseline.

**Supplemental Table 3.**
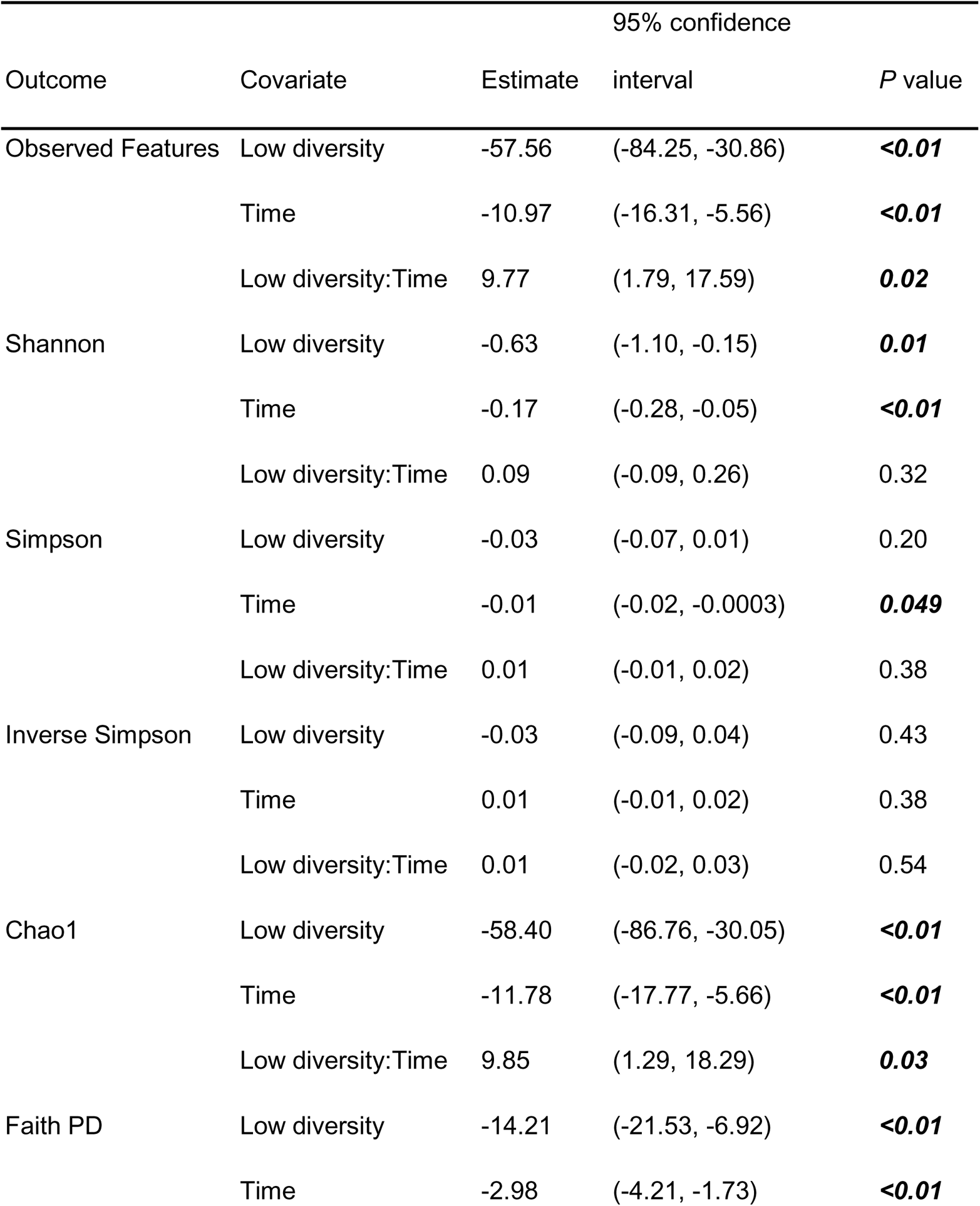
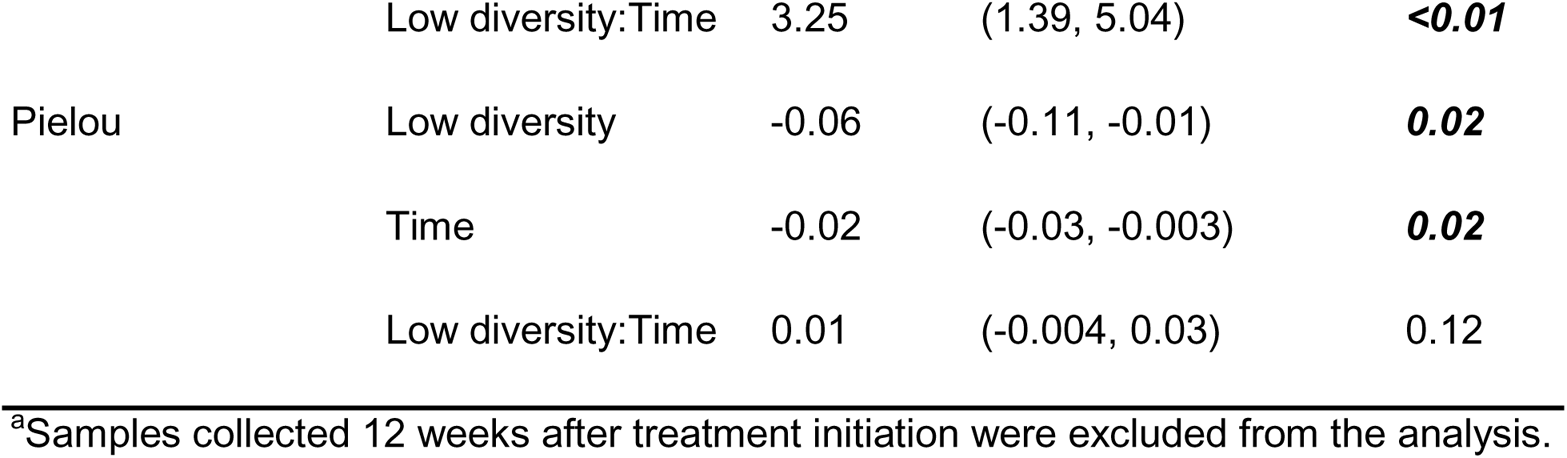
Linear Mixed Model Analysis of the Interaction Effect of Time^a^ and Baseline Diversity on Gut Diversity Throughout Treatment.

**Supplemental Table 4.**
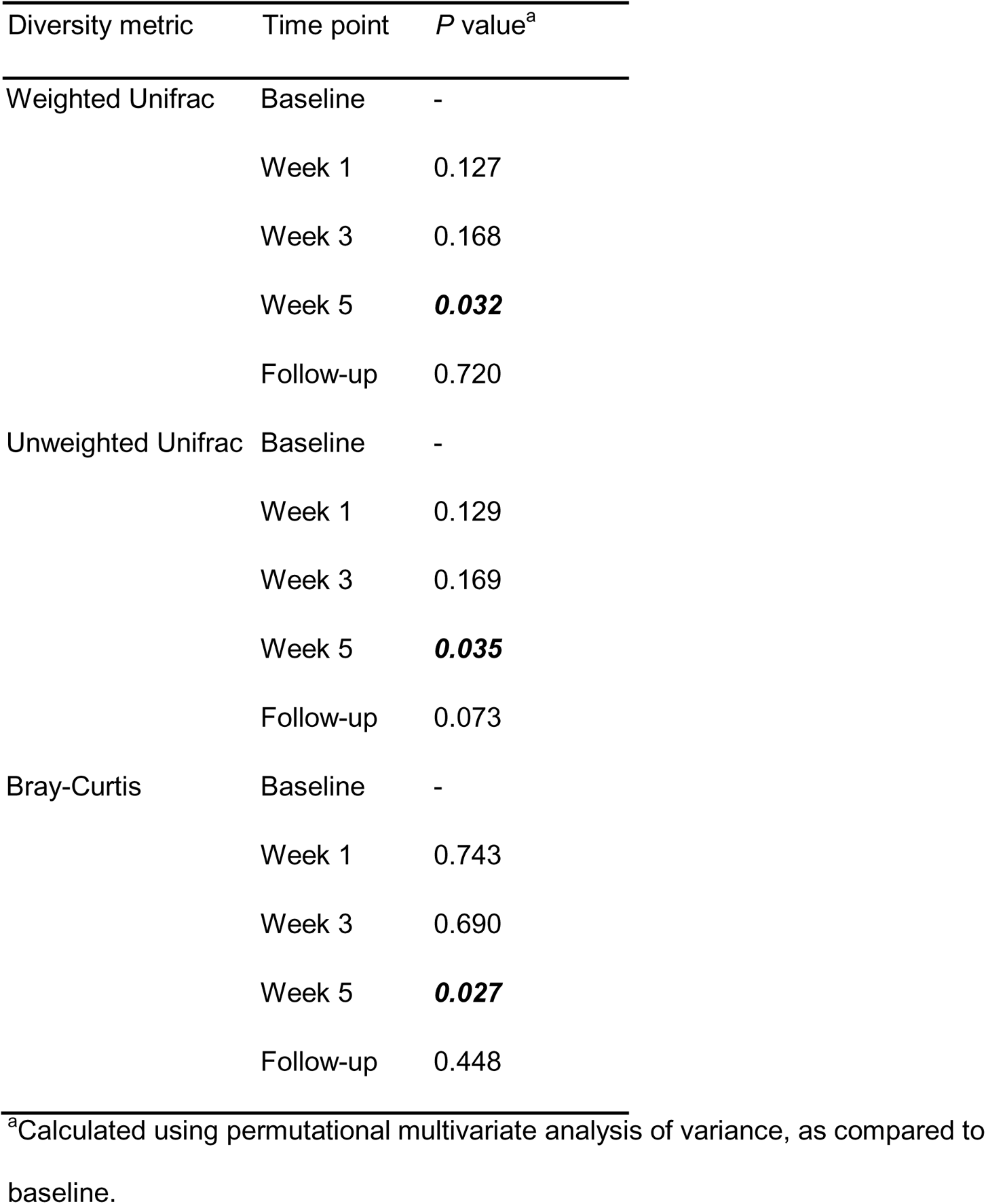
Beta Diversity Metrics at Baseline vs. Those During and After Chemoradiotherapy.

| Diversity metric | Time point | *P* value<sup>a</sup> |
| --- | --- | --- |
| Weighted Unifrac | Baseline | - |
|  | Week 1 | 0.127 |
|  | Week 3 | 0.168 |
|  | Week 5 | **0.032** |
|  | Follow-up | 0.720 |
| Unweighted Unifrac | Baseline | - |
|  | Week 1 | 0.129 |
|  | Week 3 | 0.169 |
|  | Week 5 | **0.035** |
|  | Follow-up | 0.073 |
| Bray-Curtis | Baseline | - |
|  | Week 1 | 0.743 |
|  | Week 3 | 0.690 |
|  | Week 5 | **0.027** |
|  | Follow-up | 0.448 |

<sup>a</sup>Calculated using permutational multivariate analysis of variance, as compared to baseline.

**Supplemental Table 5.** Changes in the Relative Abundances of Taxa, Baseline vs. Week 5 and Week 5 vs. Follow-Up.

| Taxa | Median (range) |  |  | P value<sup>a</sup> |  |
| --- | --- | --- | --- | --- | --- |
|  | Baseline | Week 5 | Follow-up | Baseline vs. week 5 | Week 5 vs. follow-up |
| Clostridia (class) | 0.37 (0.14-0.58) | 0.21 (0.08-0.55) | 0.41 (0.19-0.6) | **0.033** | **0.004** |
| Actinobacteria (phylum) | 0.01 (0-0.11) | 0.07 (0.01-0.48) | 0.01 (0-0.27) | **0.008** | 0.064 |
| Actinobacteria (class) | 0.01 (0-0.11) | 0.06 (0-0.48) | 0.01 (0-0.27) | **0.003** | 0.064 |
| *Corynebacteriaceae* (family) | 0 (0-0.05) | 0.05 (0-0.46) | 0 (0-0.01) | **0.002** | **0.002** |
| Corynebacteriales (order) | 0 (0-0.05) | 0.05 (0-0.46) | 0 (0-0.02) | **0.002** | **0.002** |
| *Corynebacterium* (genus) | 0 (0-0.03) | 0.04 (0-0.45) | 0 (0-0.01) | **0.001** | **0.006** |
| *Pasteurellaceae* (family) | 0 (0-0.04) | 0.01 (0-0.35) | 0 (0-0.02) | 0.065 | 0.059 |
| Pasteurellales (order) | 0 (0-0.04) | 0.01 (0-0.35) | 0 (0-0.02) | 0.065 | 0.059 |
| Lentisphaeria (class) | 0 (0-0) | 0 (0-0.002) | 0 (0-0.002) | 0.371 | 1.000 |
| Victivallales (order) | 0 (0-0) | 0 (0-0.002) | 0 (0-0.002) | 0.371 | 1.000 |
| Lentisphaerae (phylum) | 0 (0-0) | 0 (0-0.002) | 0 (0-0.002) | 0.371 | 1.000 |
| *Veillonellaceae* (family) | 0.03 (0-0.14) | 0.02 (0-0.04) | 0.02 (0-0.09) | **0.040** | 0.695 |
| Selenomonadales (order) | 0.04 (0.02-0.15) | 0.02 (0.01-0.05) | 0.03 (0-0.11) | **0.008** | 0.105 |
| Negativicutes (class) | 0.04 (0.02-0.15) | 0.02 (0.01-0.05) | 0.03 (0-0.11) | **0.008** | 0.105 |
| Clostridiales (order) | 0.37 (0.14-0.58) | 0.21 (0.08-0.55) | 0.41 (0.19-0.6) | **0.033** | **0.004** |
| Firmicutes (phylum) | 0.48 (0.25-0.62) | 0.29 (0.14-0.57) | 0.47 (0.36-0.77) | **0.005** | **0.002** |
| *Ruminococcaceae* (genus) | 0 (0-0.004) | 0 (0-0.001) | 0 (0-0) | 0.059 | 0.100 |
| *Finegoldia* (genus) | 0.01 (0-0.11) | 0 (0-0.02) | 0.02 (0-0.18) | 0.127 | **0.006** |
| Methanobacteria (class) | 0 (0-0) | 0 (0-0) | 0 (0-0) | 0.371 | 0.181 |
| *Methanobacteriaceae* (family) | 0 (0-0) | 0 (0-0) | 0 (0-0) | 0.371 | 0.181 |
| *Subdoligranulum* (genus) | 0 (0-0.09) | 0 (0-0.02) | 0.01 (0-0.07) | 0.078 | **0.033** |
| *Haemophilus* (genus) | 0 (0-0.04) | 0 (0-0.29) | 0 (0-0.01) | 0.147 | 0.151 |

<sup>a</sup>Calculated using a Wilcoxon signed-rank test.

**Supplemental Table 6.** Changes in Baseline Relative Abundances of Taxa between Patients With High or Low Gastrointestinal Toxicity.

| Taxa | High toxicity, median<br>(range) | Low toxicity, median<br>(range) | *P* value<sup>a</sup> |
| --- | --- | --- | --- |
| Actinobacteria (phylum) | 0.0046 (0.00051-0.013) | 0.018 (0.0057-0.11) | 0.052 |
| Actinobacteria (class) | 0.0042 (0.00-0.11) | 0.016 (0.0049-0.11) | **0.030** |
| Clostridia (class) | 0.33 (0.18-0.35) | 0.38 (0.29-0.50) | **0.030** |
| Clostridiales (order) | 0.33 (0.18-0.35) | 0.38 (0.29-0.50) | **0.030** |
| *Clostridiales FamilyXI* (family) | 0.10 (0.025-0.22) | 0.27 (0.11-0.33) | 0.052 |
| *Peptoniphilus* (genus) | 0.0040 (0.00-0.076) | 0.069 (0.023-0.16) | 0.052 |
| *Sellimonas* (genus) | 0.00018 (0.00-0.00044) | 0.00 (0.00,0.00) | **0.048** |

<sup>a</sup>Calculated using a Wilcoxon signed-rank test.

